# Causal Forests versus Inverse Probability of Treatment Weighting to adjust for Cluster-Level Confounding: A Parametric and Plasmode Simulation Study based on US Hosptial Electronic Health Record Data

**DOI:** 10.1101/2025.04.29.25326430

**Authors:** Mike Du, Stephen Johnston, Paul M Coplan, Victoria Y. Strauss, Sara Khalid, Daniel Prieto-Alhambra

## Abstract

**Background:** Rapid innovation and new regulations increase the need for post-marketing surveillance of implantable devices. However, complex multi-level confounding related to patient-level and surgeon or hospital covariates hampers observational studies of risks and benefits. We conducted two simulation studies to compare the performance of Causal Forests (CF) vs Inverse Probability of Treatment Weighting (IPTW) to reduce confounding bias in the presence of strong surgeon impact on treatment allocation.

**Methods:** Two Monte Carlo simulation studies were carried out: 1) Parametric simulations with patients nested in clusters (ratio 10:1, 50:1, 100:1, 200:1, 500:1) and sample size n=10,000 were conducted with patient and cluster level confounders; 2) Plasmode simulations generated from a cohort of 9,981 patients admitted for pancreatectomy between 2015 to 2019 from the US PINC AT™ hospital research database. Different CF algorithms and IPTW were used to estimate binary treatment effects.

**Results:** CF provided more accurate estimates when the cluster-level confounding effect was strong (OR=2.5): relative bias 11.2% (11.77, 11.76) for CF compared with 19.9% (19.26, 20.54) for IPTW.

**Conclusions:** CF shows promise as a method for estimating treatment effects in scenarios where cluster-level confounding strongly impacts treatment allocation. More research is needed to guide its use.

**Key messages:**

- Causal Forests using the double regression tree algorithm showed the least bias in scenarios with strong cluster-level confounding and small cluster size.
- Including the cluster-ID indicator in Causal Forests resulted in higher bias and empirical standard error in scenarios with fewer but larger size clusters.
- Causal Forests outperformed IPTW in scenarios with strong cluster-level confounding.

**Plain language summary:** This study compared two methods for estimating treatment effects in clustered observational health data where the healthcare provider, such as a hospital or surgeon, influences patient outcomes and treatment allocation. Using simulated data, we evaluated a machine learning method called Causal Forest and compared it to a commonly used approach called Inverse Probability of Treatment Weighting (IPTW). We found that Causal Forest, particularly when using the double regression tree technique, performed well in many scenarios and gave less bias when provider-related factors strongly influenced the treatment allocation. While IPTW generally produces accurate results, it depends on strong assumptions that may not hold in real-world studies. Our findings suggest that Causal Forest is a promising approach for estimating treatment effects in clustered observational health data settings, particularly in surgical and medical device research where treatment decisions vary by provider.

## Introduction

Observational studies using routinely collected patient data, such as data from health registries, hospitals, or insurers, have become essential for clinical treatment comparative studies, especially when randomised controlled trials (RCTs) are unfeasible or unethical (1). In surgical and medical device epidemiology, treatment allocation is not only influenced by patient characteristics but also significantly by hospital and physician/surgeon characteristics, leading to clustered data. This introduces confounding by indication that is more complex than general pharmacoepidemiology studies (2). The influence of surgeons or providers can heavily bias treatment effect estimates, presenting a unique challenge in accurately assessing the treatment effects of surgical procedures or medical devices. Rosenbaum and Rubin (3, 4) introduced the propensity score (PS) to adjust for confounding, and while PS-based methods are popular in pharmacoepidemiology, they were initially designed without taking the clustered nature of data due to surgeon or hospital characteristics in surgical and device studies and can often fail to account for cluster-level confounding, where the impact of cluster-level confounders is strong (5).

Recent advancements in machine learning techniques offer alternate solutions to these challenges, and the performance has been evaluated against clustered data in several recent studies (6-9). Among these, Causal Forests, a random Forest-based algorithm, has emerged as a promising method for estimating treatment effects in observational studies (10). The Causal Forest algorithm modifies the traditional Random Forest algorithm (11) for treatment effect estimation.

Causal Forests, a machine learning method for estimating treatment effects, are based on decision trees known as Causal Trees, first proposed by Wager et al (7, 8). The algorithm begins by dividing the full sample into two subsamples and applying bootstrap aggregation (11). Each subsample is then split into two parts: one for determining tree splits and the other for estimating treatment effects. This double-sample splitting reduces bias by separating the tree split determination from treatment effect estimation, though it may result in lower precision due to reduced data usage. The trees continue splitting until further divisions no longer improve the heterogeneity of treatment effects or predefined criteria, such as minimum leaf size or maximum depth, are met. The final treatment effect is calculated as a weighted average of the effects from each tree (7). While double-sample trees reduce bias, they discard proportions of the sample, limiting precision. To address this, propensity scores can be used to guide splits, though misspecified propensity scores may introduce bias as with other propensity score-based methods (5, 12).

While several studies have compared Causal Forests with propensity score-based methods and demonstrated that Causal Forests shown accuracy similar to those of propensity score methods (10, 13), limited attention has been given to the performance of Causal Forests in surgical epidemiology settings. In these settings, cluster-level covariates may significantly impact treatment allocation, potentially influencing the accuracy of treatment effect estimates. Therefore, we aimed to compare the precision and bias of Causal Forests with Inverse Probability of Treatment Weighting (IPTW), a method that has been shown to be less accurate in scenarios with strong cluster-level confounders impact on treatment allocation from a previous study we conducted (14). Our study focuses on various scenarios with differing degrees of cluster-level confounding on treatment allocation. To evaluate this, we employed parametric and plasmode simulations (15, 16), in which covariates were simulated based on a real-world dataset.

## Methods

### Parametric Simulation data generation process

The simulation settings were based on previous simulation studies with clustered data (17, 18), but with parameters chosen to mimic the structure of a real-world dataset described below. We simulated the datasets via parametric simulations (19-21) with a fixed sample size of 10,000 individuals to represent the patients, binary treatment allocation (T), and binary outcome (Y). The datasets contain seven patient-level covariates (x1 to x7), two cluster-level covariates (z1 and z2 to represent potential hospital-level or surgeon-level confounders), and a cross-level interaction term between the individual and cluster-level confounders, which were simulated for each patient. Among the individual covariates simulated, 5 were confounders (x1-x5), one (x6) was an instrumental variable associated with the treatment but not with the outcome (other than through the treatment), and x7 was a risk factor associated with the outcome but not the treatment (22). Both cluster-level covariates (z1 and z2) were generated as confounders associated with treatment and outcome. The cluster and patient-level covariates were simulated from different probability distributions to reflect different covariates observed in real-world medical devices or surgical data.

20 different scenarios were simulated to test the performance of the proposed methods for controlling cluster-level confounding. The scenarios were generated by varying the cluster structure of the data and the effect size of the cluster level confounders on treatment allocation (z1 and z2), ranging from negligible with odds ratio =1.01 to odds ratio = 2.5 to resemble strong cluster level confounding. Five different cluster structures were simulated with different cluster numbers (m) and average patients per cluster (n) (m=10, n = 1000), (m=50, n=200), (m=100, n=100), (m=200, n=50) and (m=500, n=20). Patients per cluster (n) were randomly sampled from the Poisson distribution with mean n for each cluster within the dataset. Table 1 gives the 20 different simulation data scenarios generated. Figure 1 gives the Causal diagram of the simulation covariates and the simulations are run for 100 repetitions.

**Table 1:**
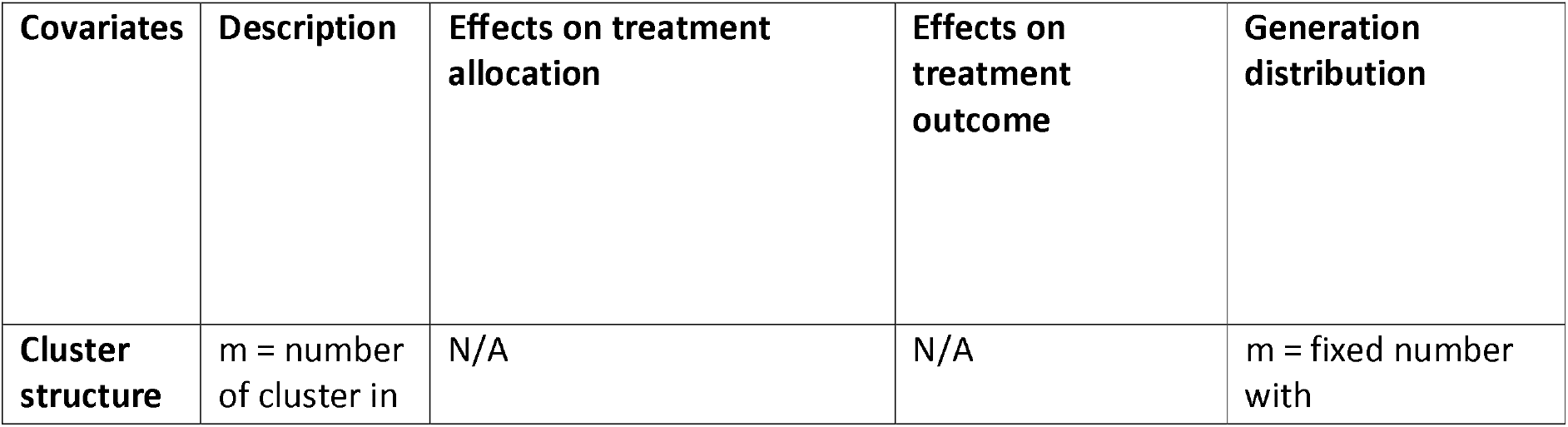
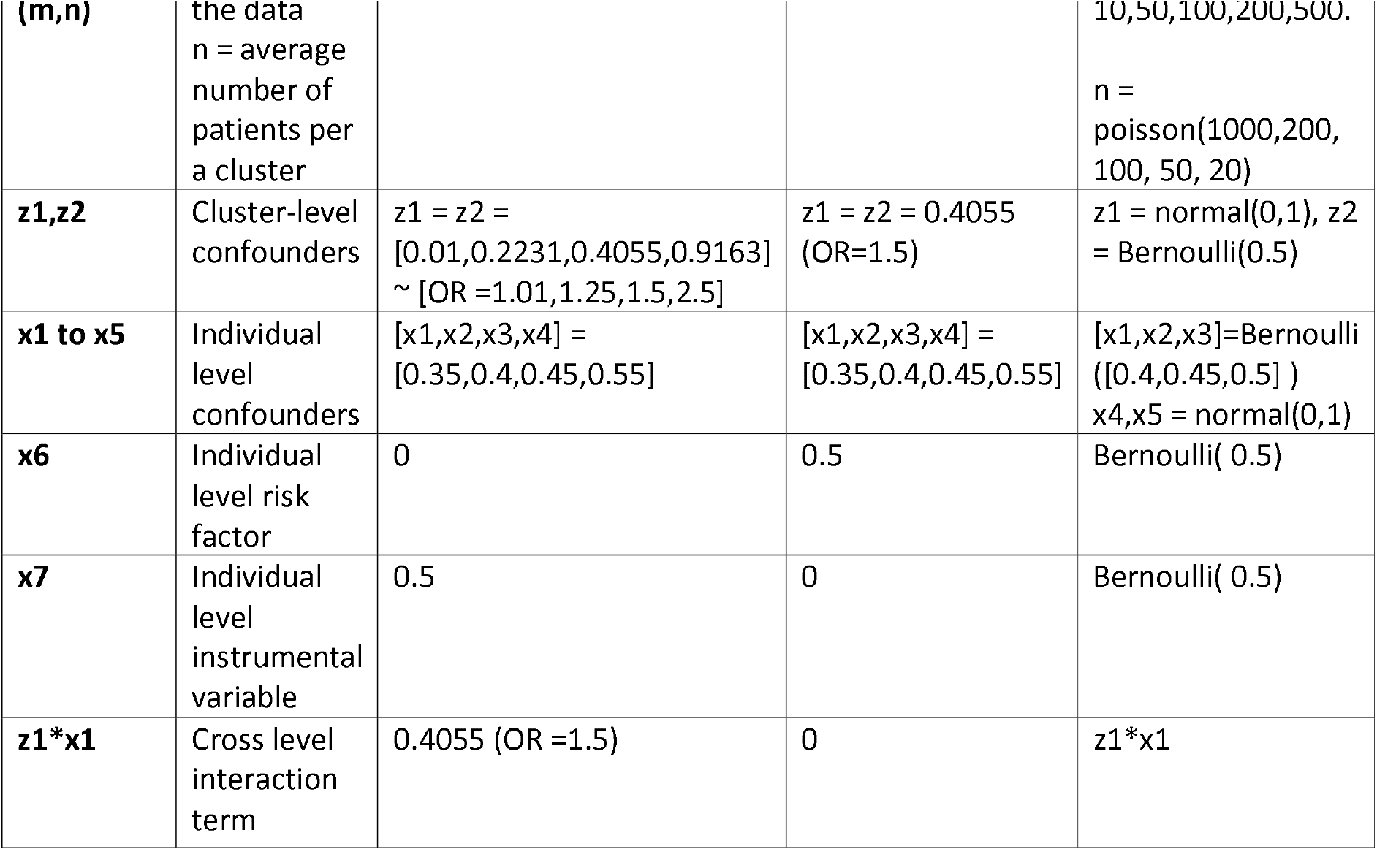
The table gives the generation distribution, effects on treatment allocation and effects on treatment outcome for covariates generated in the simulations. OR = odd ratio.

| Covariates | Description | Effects on treatment allocation | Effects on treatment outcome | Generation distribution |
| --- | --- | --- | --- | --- |
| Cluster structure | m = number of cluster in | N/A | N/A | m = fixed number with |
| **(m,n)** | the data<br>n = average<br>number of<br>patients per<br>a cluster |  |  | 10,50,100,200,500.<br>n =<br>poisson(1000,200,<br>100, 50, 20) |
| **z1,z2** | Cluster-level<br>confounders | z1 = z2 =<br>[0.01,0.2231,0.4055,0.9163]<br>~ [OR =1.01,1.25,1.5,2.5] | z1 = z2 = 0.4055<br>(OR=1.5) | z1 = normal(0,1), z2<br>= Bernoulli(0.5) |
| **x1 to x5** | Individual<br>level<br>confounders | [x1,x2,x3,x4] =<br>[0.35,0.4,0.45,0.55] | [x1,x2,x3,x4] =<br>[0.35,0.4,0.45,0.55] | [x1,x2,x3]=Bernoulli<br>([0.4,0.45,0.5] )<br>x4,x5 = normal(0,1) |
| **x6** | Individual<br>level risk<br>factor | 0 | 0.5 | Bernoulli( 0.5) |
| **x7** | Individual<br>level<br>instrumental<br>variable | 0.5 | 0 | Bernoulli( 0.5) |
| **z1\*x1** | Cross level<br>interaction<br>term | 0.4055 (OR =1.5) | 0 | z1*x1 |

**Figure 1:**
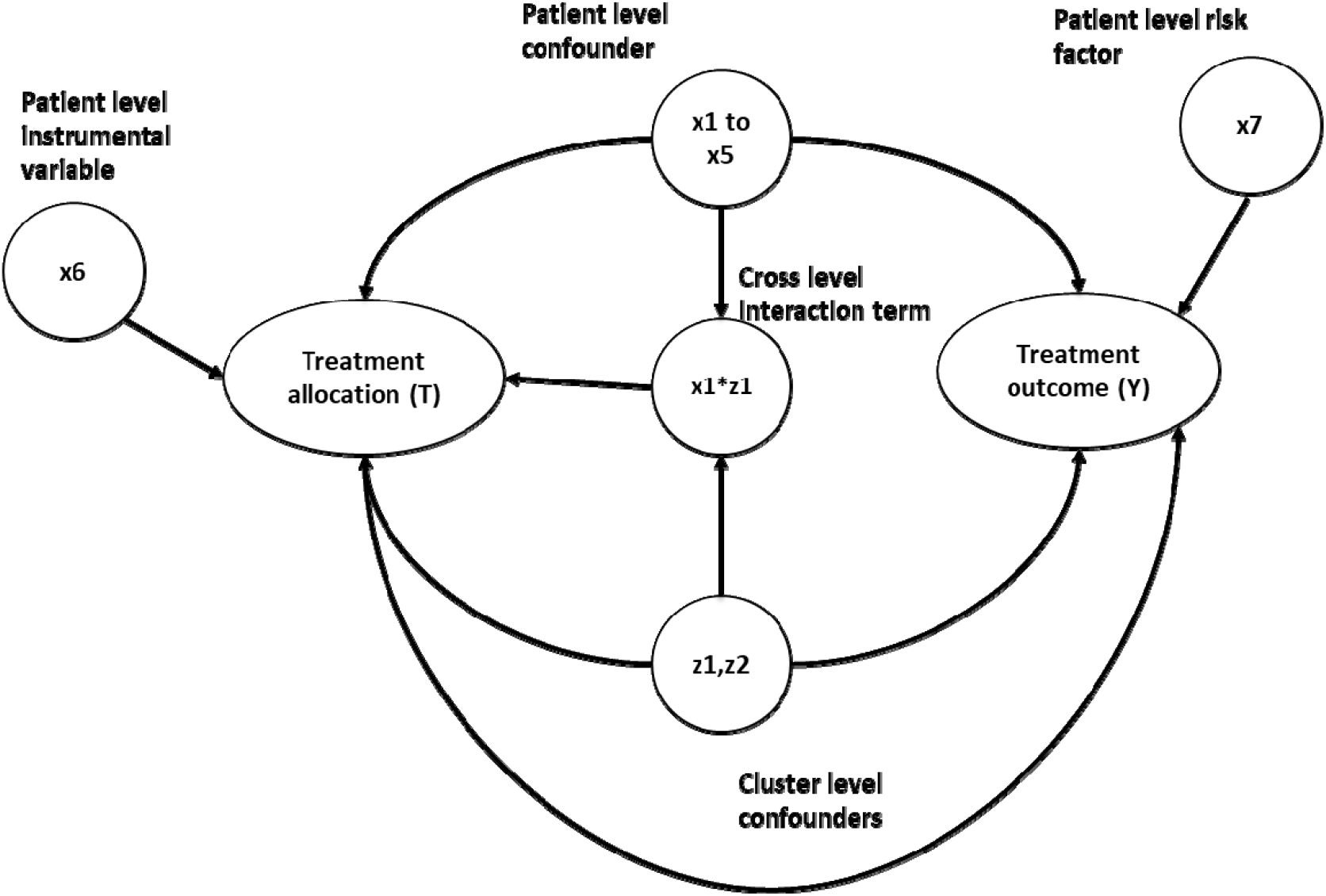
This diagram gives the Causal relationship between the covariates in the simulation data, the arrow indicates causes. For example, x1-> Y implies x1 causes Y.

### Plasmode simulation data generation process

Plasmode simulation (16) is a method that generates synthetic data by re-sampling from pre-selected observed covariates of a real-world dataset. The re-sampling of covariates was performed using the bootstrap with replacement method (23). The exposure and outcome of Plasmode simulation data are generated using the investigators’ pre-specified re-sampled covariates from the real-world data cohort and choice of true treatment effects. Hence, simulation data generated using Plasmode simulation will preserve the data structure and covariates of the real-world data cohort from which it generated the simulated data. The covariates in Plasmode simulation are more closely matched to real-world data than the Parametric simulation. However, Plasmode simulation lacks the ability to change its data structure.

The real-world data cohort we generated in our Plasmode simulation were from the US PINC AI™ Healthcare Database, an all-payer hospital database collected from among over 1000 hospitals in the US (24). The Premier Healthcare Database includes information from hospitals’ electronic health records, including diagnoses, procedures, patient characteristics, and hospital features. The cohort included 9981 patients aged 18 or over that were admitted for pancreatectomy from 2015-2019. The patients’ covariates of the simulated data were generated based on age, sex, and the Charlson comorbidity index. The cluster of the simulated data was identified using the hospital ID. The cohort had 341 unique hospital IDs, with an average of 30 patients per hospital. The cluster covariates were re-sampled from hospital-related covariates such as type (teaching or not teaching), hospital size (500+ or 500 fewer beds), and the hospital’s yearly pancreatectomy volumes etc. A full summary of the covariates used in the Plasmode simulation is provided in the supplementary materials (s1 and s2).

### Inverse Probability of treatment weighting

For IPTW, the propensity scores were estimated using a logistic regression propensity score model with cluster-level and patient-level confounders as covariates. Then, the average treatment effects were estimated using a random effects model. The treatment outcome was regressed on the treatment allocation using a random intercept logit model weighted with stabilised IPTW calculated from the PS model. The method used is similar to the IPTW methods used in previous methodology research for clustered data, which has shown to be effective in reducing bias due to cluster-level confounding (25).

### Causal Forests

For Causal Forests, three different methods were implemented by changing either the input covariates or the tuning parameters of the Causal Forest. 1), the treatment estimate was computed using the double regression trees method as explained in the Introduction section. 2), the double regression tree method was used, same as (1), but with the cluster label added as input for the Causal Forests. This meant binary dummy covariates for each cluster to indicate which cluster this patient belongs to. 3), the propensity score was used as decision parameters for splitting, as explained in the introduction. Table 2 of all the methods compared in this study. The Causal Forest was implemented using the grf package in R version 2.2.0 (26).

**Table 2:**
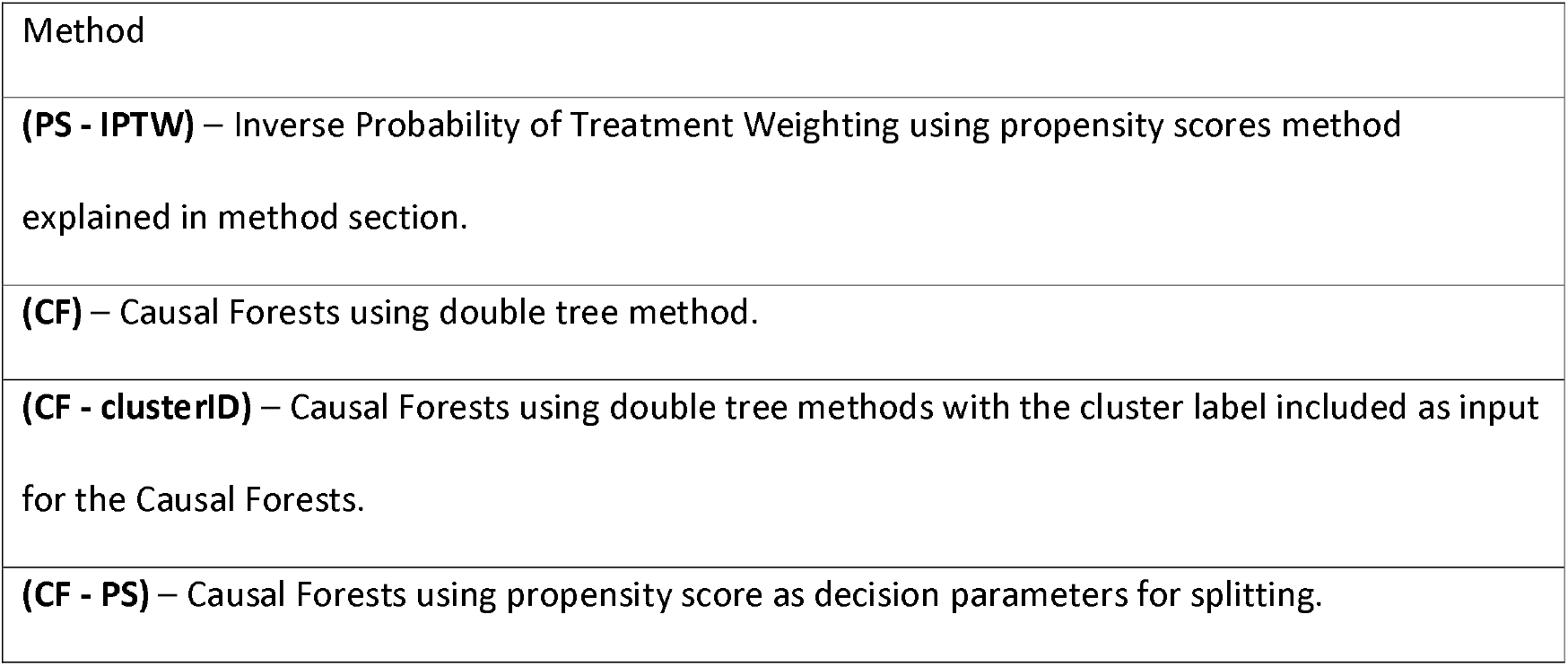
Methods compared in this study.

**(PS - IPTW)** – Inverse Probability of Treatment Weighting using propensity scores method explained in method section.

**(CF)** – Causal Forests using double tree method.

**(CF - clusterID)** – Causal Forests using double tree methods with the cluster label included as input for the Causal Forests.

**(CF - PS)** – Causal Forests using propensity score as decision parameters for splitting.

### Assessment of results

The precision and accuracy of the estimated treatment effects for both propensity score weighting and Causal Forests were compared. Using average absolute relative bias (Rbias), empirical standard error (EmpSE) and 95% confidence intervals model coverage (95% Coverage) of the 1000 repetitions for each simulated scenarios as defined in the guidance literature on simulation studies by Morris et al (27). All analyses were performed in R version 4.3.1, with the Parametric simulation data generated with the “simstudy” package (28) and the Plasmode simulation data generated with the “Plasmode” package (16).

## Results

The findings from the Parametric simulations, shown in figures 2 – 5, indicate that CF using the double regression tree approach performed the best out of the three different method tested in this study. CF gave the lowest relative bias and empirical standard error compared to CF-PS. For example, in the cluster structure scenario with m=10 and n=1000 with cluster-level confounders effect on treatment allocation OR=1.5, CF had a relative bias of 13.8%. In comparison, CF-PS had a relative bias of 22.2%. These results suggest that incorporating propensity scores as the decision parameter for splitting within the Causal Forest did not improve the accuracy and precision of the treatment estimate for the clustered data scenarios tested in this simulation study.

**Figure 2:**
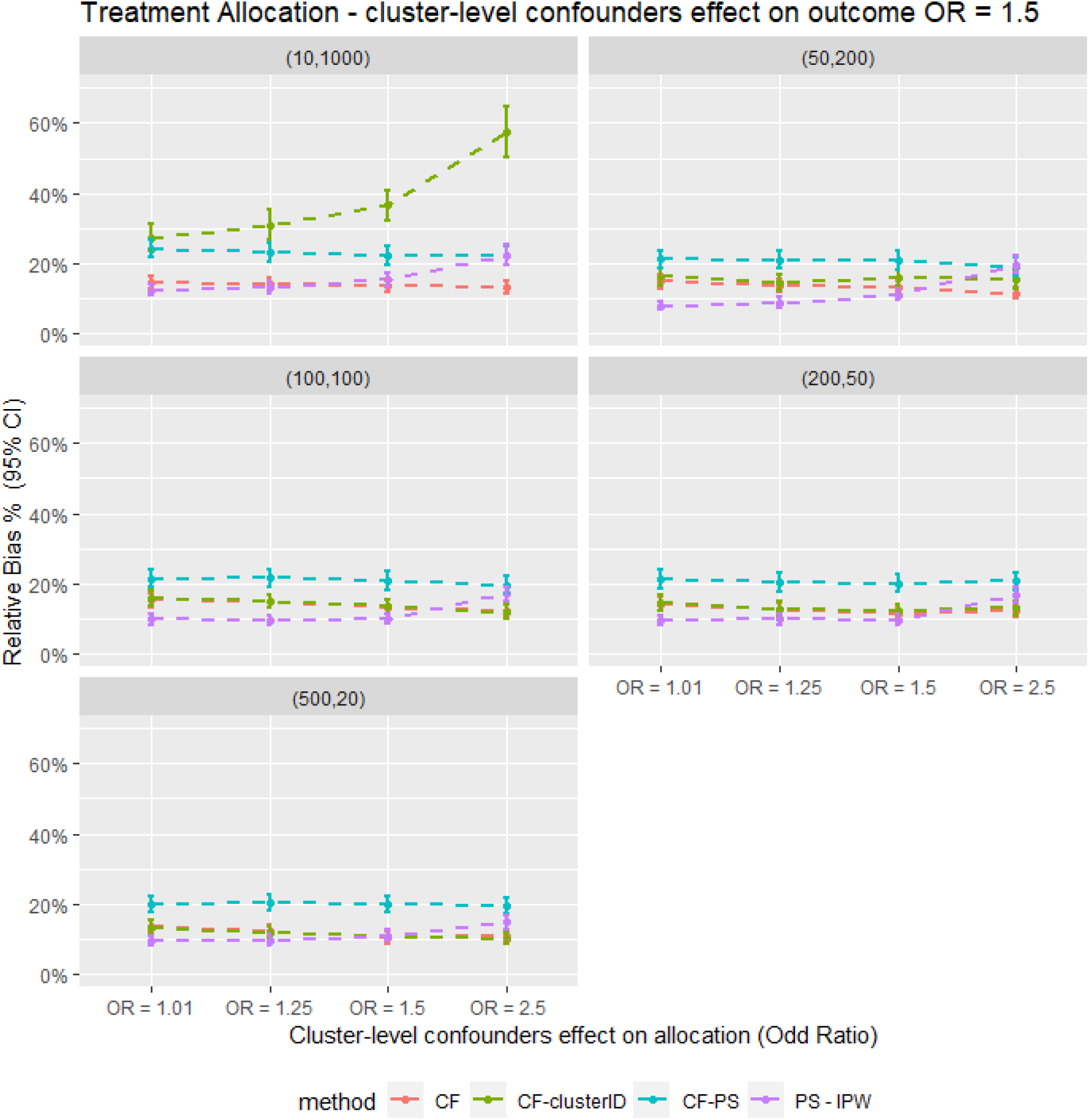
Average relative bias for different confounder effect on treatment allocation scenarios for different methods for all the data scenario tested in the Parametric simulation study. PS - IPTW – Inverse Probability of Treatment Weighting using propensity scores method, explained in method section, CF – Causal Forests using double tree method. CF - clusterID – Causal Forests using double tree methods with the cluster label included as input for the Causal Forests, CF - PS – Causal Forests using propensity score as decision parameters for splitting., (XX,XX) = cluster structure with (number of cluster, average patients per cluster).

Moreover, including the cluster ID indicator as input for the Causal Forest (CF-clusterID) did not improve the accuracy of CF. Instead, it substantially decreased the accuracy of CF in the cluster structure scenarios with m=10 and n=1000. For example, the relative bias for CF was 14.0%, compared with 30.9% for CF-clusterID in the cluster-level confounder effects on treatment allocation (OR=1.25) scenario. However, as the number of clusters in the structure increased and the number of individuals per cluster decreased, the differences in relative bias between CF and CF-clusterID also decreased. Nevertheless, CF still had slightly lower relative bias and empirical standard error in the scenarios tested. These findings suggest that including the cluster ID as input for Causal Forest is unnecessary for data with cluster-level confounding, regardless of the cluster structure of the data.

Additionally, the Parametric simulation results indicated that IPTW outperformed CF in most scenarios. For instance, in scenarios with a cluster structure of m=50 and n=200 and cluster-level confounding on treatment allocation (OR=1.25), the relative bias was 8.93% for IPTW compared to 13.7% for CF. However, CF provided more accurate estimates when the cluster-level confounding effect was strong (OR=2.5). For example, in scenarios with a cluster structure of m=500 and n=20 and cluster-level confounding effect on allocation of OR=2.5, the relative bias was 11.1% for CF compared with 19.7% for IPTW. These results suggest that CF could be a suitable alternative to IPTW in scenarios with strong cluster-level confounders effect on treatment allocation because it gives results with lower bias under such conditions.

Furthermore, as shown in figure 3 the parametric simulations showed that the empirical standard error was similar for all the methods compared in this study, except for CF-clusterID, which had substantially higher standard error in scenarios with m=10 and n=1000. This suggests that the precision of Causal Forest based methods were comparable to that of IPTW. Consequently, the trends observed in term of 95% CI model coverage (figure 4) were similar to those observed for the relative bias. In scenarios with a strong cluster-level confounding effect on allocation (OR = 2.5), CF offered higher model coverage. Conversely, in other scenarios, IPTW gave higher model coverage than CF.

**Figure 3:**
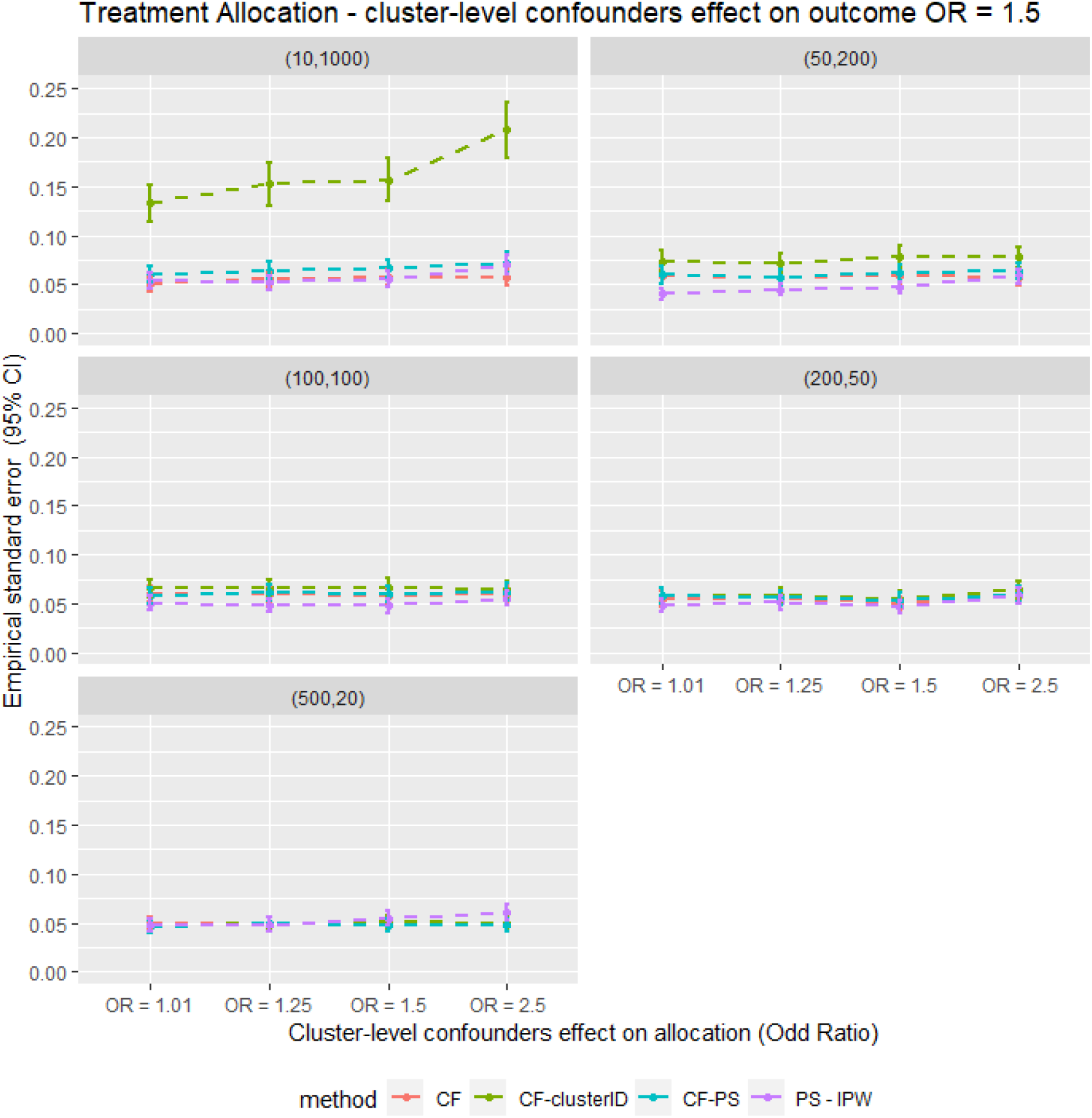
Average empirical standard error for different confounder effect on treatment allocation scenarios for different methods for all the data scenario tested in the Parametric simulation study. PS - IPTW – Inverse Probability of Treatment Weighting using propensity scores method, explained in method section, CF – Causal Forests using double tree method. CF - clusterID – Causal Forests using double tree methods with the cluster label included as input for the Causal Forests, CF - PS – Causal Forests using propensity score as decision parameters for splitting., (XX,XX) = cluster structure with (number of cluster, average patients per cluster).

**Figure 4:**
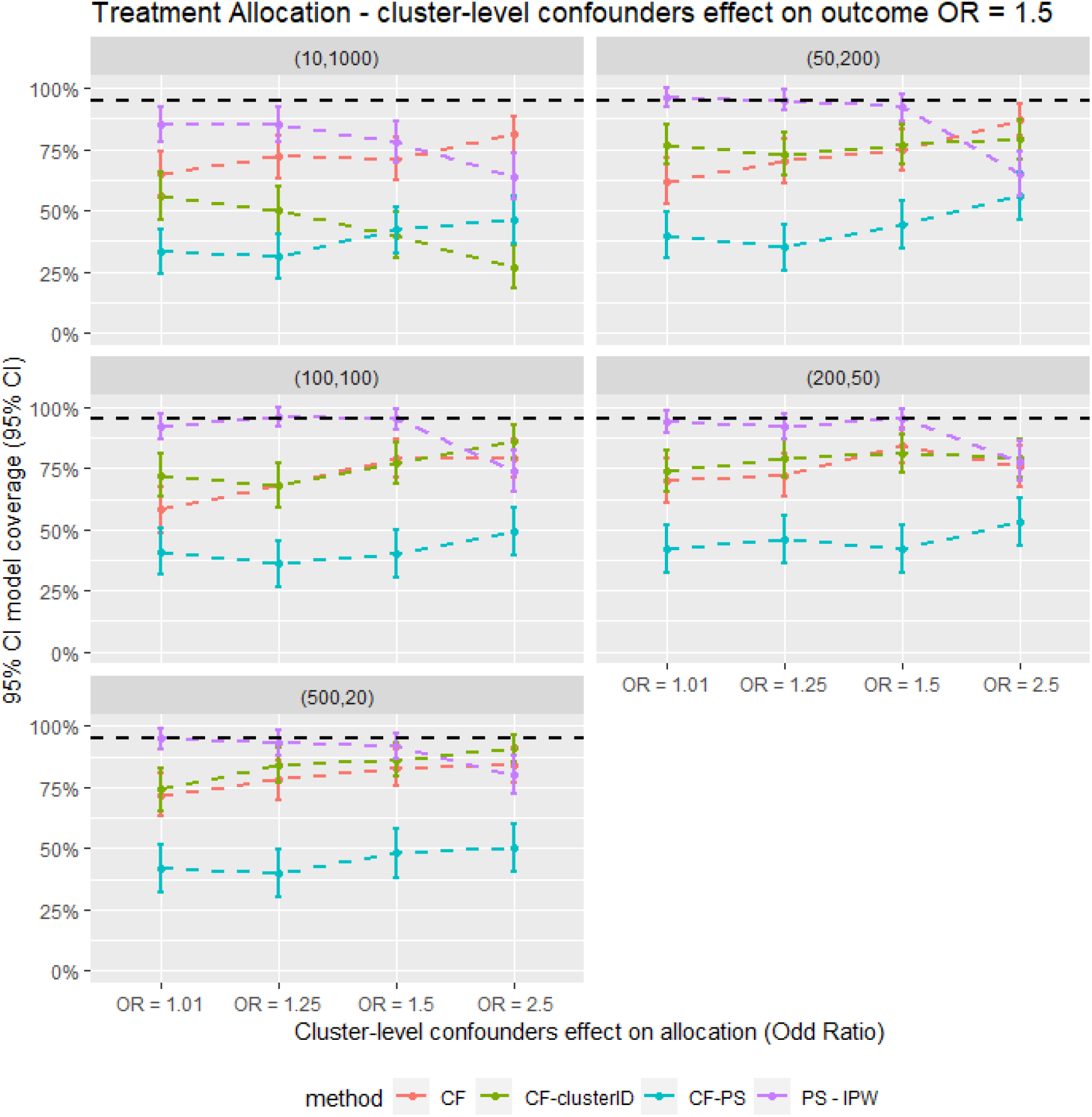
Average model coverage for different confounder effect on treatment allocation scenarios for different methods for all the data scenario tested in the Parametric simulation study. PS - IPTW – Inverse Probability of Treatment Weighting using propensity scores method, explained in method section, CF – Causal Forests using double tree method. CF - clusterID – Causal Forests using double tree methods with the cluster label included as input for the Causal Forests, CF - PS – Causal Forests using propensity score as decision parameters for splitting., (XX,XX) = cluster structure with (number of cluster, average patients per cluster).

The Plasmode simulation results (figure 5) also show that the performance of the CF and PS-IPTW methods in estimating treatment effects are comparable to those of the parametric simulation, which had a cluster structure with m=500 and n=20. Specifically, the results for CF and CF-clusterID were almost identical, indicating that adding the cluster ID indicator does not affect treatment effect estimates in data with a large number of clusters and small cluster sizes. The PS-IPTW method showed slightly better accuracy in treatment estimates compared to CF, with lower relative bias, regardless of the strength of the cluster effect on treatment allocation. However, the 95% confidence intervals of the relative bias for both methods overlapped, and the model coverage of the Plasmode simulation was also similar between the two methods. This suggests that the improvement in accuracy of PS-IPTW over CF was minimal, consistent with the findings from the parametric simulation.

**Figure 5:**
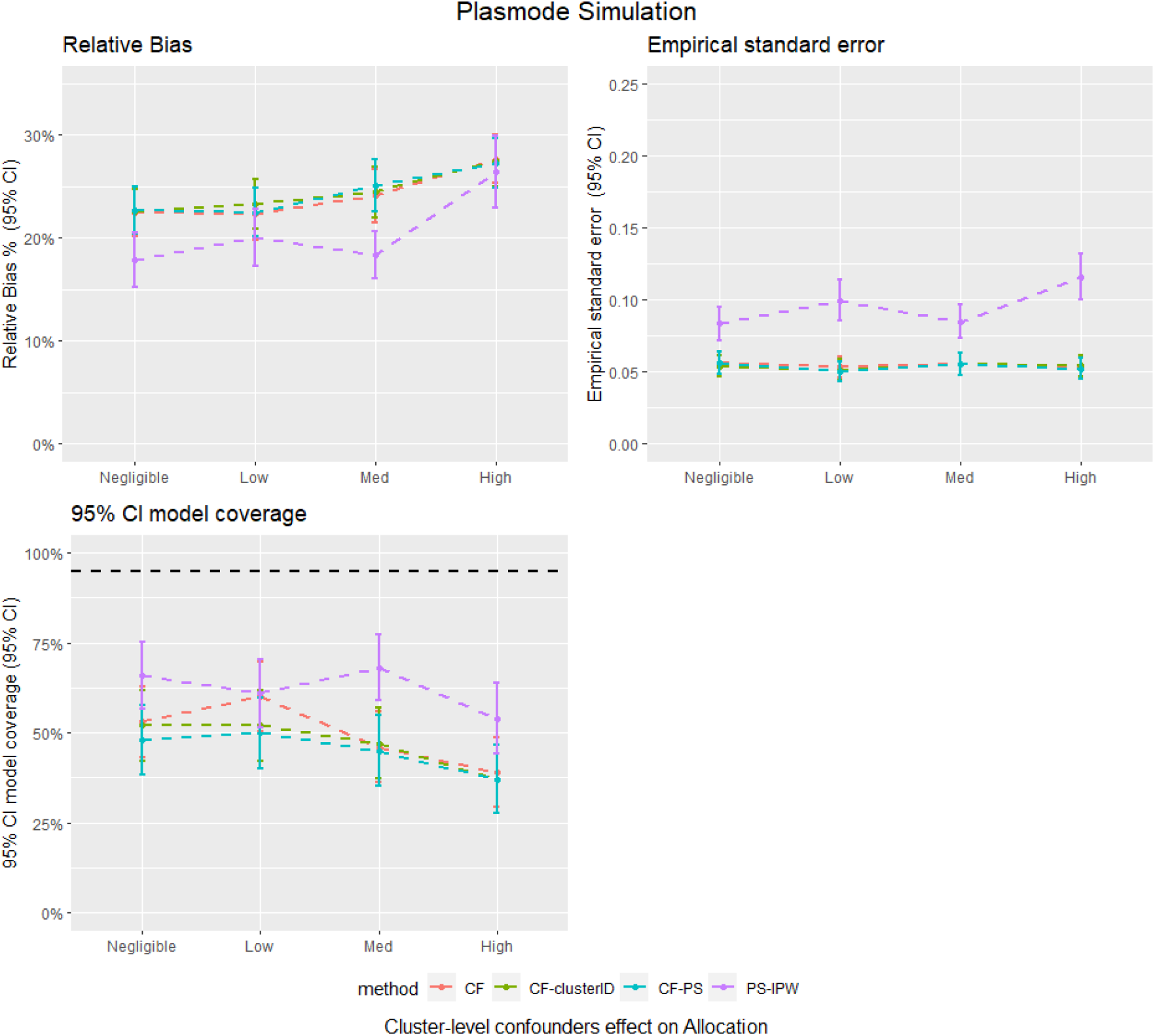
Average relative bias, model coverage and empirical stand error for different matching strategy for all the data scenario tested in the Plasmode simulation study. PS - IPTW – Inverse Probability of Treatment Weighting using propensity scores method, explained in method section, CF – Causal Forests using double tree method. CF - clusterID – Causal Forests using double tree methods with the cluster label included as input for the Causal Forests, CF - PS – Causal Forests using propensity score as decision parameters for splitting.

However, there were several trends observed in the Plasmode simulation study that were different to the Parametric simulation study. Firstly, CF-PS had similar results to CF in terms of relative bias and model coverage in the Plasmode simulation, whereas in the parametric simulation, CF-PS had higher bias and lower model coverage than CF. Additionally, the empirical standard error for PS-IPTW was higher than that of the Causal Forest-based method tested in the Plasmode simulation, whereas in the parametric simulation, the empirical standard error was similar for both methods. Finally, the relative bias for the Causal Forest-based method increased as the cluster-level confounder effect on treatment allocation increased in the Plasmode simulation, but not in the parametric study.

## Discussion

This study reveals several important findings regarding the performance of Causal Forests compared to IPTW. First, the results indicated Causal Forest using the double regression tree algorithm produced the most favourable performance in terms of bias. Then, incorporating the cluster-ID indicator in the Causal Forest model did not have a noticeable effect on the accuracy or precision of the treatment effects in majority of the simulation settings tested. However, in situations where the number of clusters was small and the cluster size was large, the addition of the cluster-ID indicator resulted in significantly higher bias and empirical standard error when compared to Causal Forest methods without this indicator. Furthermore, the inclusion of propensity score did not improve the performance of Causal Forest, as evidenced by slightly higher bias treatment estimates in the parametric simulation study and no reduction in bias in the Plasmode simulation, when compared with Causal Forest using double regression tree. These findings align with those reported in a prior simulation study conducted by Suk et al (10). Hence according to the findings of this simulation study, Causal Forest using the double regression tree method without the inclusion of the cluster-ID indicator is the most effective of the three Causal Forest models for estimation of treatment effect for clustered data with confounder from cluster-level.

We also found that when cluster-level confounding effects on treatment allocation are strong, Causal Forest may offer more accurate treatment effect estimates than IPTW. Specifically, in scenarios where cluster-level confounding effects were strong, the bias for IPTW tended to increase, while the bias for Causal Forest did not increase as much, particularly in the parametric simulation. However, in general, IPTW tended to provide more accurate treatment effect estimates than Causal Forest. It is important to note that when interpreting IPTW results, the assumptions of no misspecification of confounders in the propensity score model must be considered. In real-world data analysis, these assumptions are unlikely to be fully met (17, 29, 30). In contrast, Causal Forest selects covariates to include in the model and does not require prior user information on covariate type, making it more likely that the accuracy of Causal Forest estimates will hold when applied to real-world data.

However, we found some discrepancies in the findings between the Plasmode and parametric simulations. This could be caused by the differences in the data used in for the simulation. For instance, the Plasmode simulations included a larger number of covariates at both the individual and cluster levels, while the cluster structure of the data was different between the two simulations. Due to these differences in the data, it is difficult to determine the exact causes of the differences in the results between the Plasmode and parametric simulations without further investigation with extra scenarios simulated.

## Strengths and limitations

This study’s main strength is its use of simulated data, where the actual treatment effect was known. Hence using simulation studies allowed us to calculate the bias and empirical standard error for the treatment effects estimated using different methods. Therefore, the accuracy and precision of different methods can be compared. Using simulated data also allowed us to create different scenarios by varying data variables to see how each PS method behaves in different scenarios. This is usually difficult to achieve in real-world data analysis.

This study was also subject to limitations. A major limitation of this study is that it has not captured all cluster structure, cluster-confounding effect sizes and covariates scenarios that may occur in real-world data. Hence, the findings may only be generalisable to the scenarios tested in this simulation study. Besides the limitation of the simulation setting, this study did not evaluate the tuning parameters for Causal Forests such as the proportions of sample used for the split in double regression tree algorithm, tree depths and different stopping criteria, as currently there is limited guidance on tuning Causal Forests in epidemiology research (13). It is fair to argue that better performance can be achieved by experimenting with different tuning parameters for Causal Forests.

## Conclusion

This study provides valuable insights into the use of Causal Forests for estimating treatment effects in clustered data. The results demonstrated that the performance of Causal Forests was comparable to IPTW, with Causal Forests showing higher accuracy in scenarios where cluster-level confounders strongly influenced treatment allocation. This suggests that Causal Forests could potentially serve as an alternative method for analysing treatment effects in medical device and surgical epidemiology, particularly in scenarios where surgeons have a significant impact on treatment allocation.

However, further empirical research is needed to offer more guidance and insight on the use of Causal Forest in real-world studies. Since Prior research on the application of Causal Forest for estimating treatment effects in clustered data is limited. It would be useful to investigate how Causal Forest behaves in other sample sizes, cluster-confounding effects on outcome, its ability to handle unmeasured confounders, missing data, and interactions.

In conclusion, while Causal Forests show promise as a method for estimating treatment effects based on observational clustered data, further research is needed to fully understand their strengths and limitations. The results of this study provide a useful starting point and motivation for future research into the applications of Causal Forest methods for causal inference in observational study of surgical epidemiology and medical device research.

## Data Availability

The simulated data are avaliable upon requests

## Declarations

### Ethics approval and consent to participate

Not Applicable

### Consent for publication

Not Applicable

### Availability of data and materials

The study code for the study are available on GitHub. Please for access.

### Authors’ contributions

DP and MD led the conceptualisation of the study with contributions from SJ, SK, VS, PC and SJ. MD developed the code for statistical analyses and simulations. MD wrote the first draft of the manuscript. All authors read, contributed to, and approved the last version of the manuscript.

## Acknowledgements

Not Applicable

