## supplemental for "Causal Forests versus Inverse Probability of Treatment Weighting to adjust for Cluster-Level Confounding: A Parametric and Plasmode Simulation Study based on US Hosptial Electronic Health Record Data"

**Supplementary material**

Figure s1: This table presents the variables used in the plasmode simulation, along with their beta values for treatment outcome and allocation.

| Resampled Covariates | Covariates description | Variable Type (Hospital level/Patient level) | Beta value to treatment allocation | Beta value to treatment outcome |
| --- | --- | --- | --- | --- |
| Hospital location: urban/rural | Binary indicator | Hospital level | Varied depends on simulated scenarios | Varied depends on simulated scenarios |
| Hospital yearly surgery volume | Continuous covariates | Hospital level | Varied depends on simulated scenarios | Varied depends on simulated scenarios |
| Hospital size: beds 500+/ 500> | Binary indicator on number of beds | Hospital level | Varied depends on simulated scenarios | Varied depends on simulated scenarios |
| Hospital region:  Midwest  Northeast  South  West | Binary indicator for each region | Hospital level | Varied depends on simulated scenarios | Varied depends on simulated scenarios |
| Gender: Male | Binary indicator | Patient-level | 0.0059 | 0.0446 |
| Charlson comorbidity index:  cci01_mi cci02_chf cci03_pvd  cci04_cvd cci05_dem cci06_cpd cci07_ra  cci08_ulcer cci09_liv cci10_diab cci11_diabc  cci12_para cci13_renal cci14_can cci15_livc  cci16_can2 cci17_hi | Binary indicator (please refer to ICD website for definition)(32) | Patient-level | = [ -0.5377,  0.1663,  0.2085,  -0.3956,  0.3931,  0.4199,  -0.9809,  0.4428,  0.4518,  0.1991,  -0.1402,  0.2915,  -0.0850,  0.2536,  -1.1945,  -0.7572,  -0.9668] | =[0,  0,  0,  0,  0.6257,  -0.3543,  0.3292,  1.3924,  -0.7073,  0.0427,  0.2751,  -1.1135,  0.1841,  -0.2704,  0.4792,  1.0809,  1.7748] |
| Elixhauser Comorbidity Index:  elx01_chf elx02_arrhy  elx03_valv elx04_pcd elx05_pvd elx06_htn  elx07_htnc elx08_para elx09_neur elx10_cpd  elx11_diab elx12_diabc elx13_thyr elx14_renal  elx15_liv elx16_ulcer elx17_hiv elx18_lym  elx19_can2 elx20_can elx21_ra elx22_coag  elx23_obe elx24_wtl elx25_fed elx26_bla  elx27_dfa elx28_alc elx29_dabu elx30_psy  elx31_depr | Binary indicator (please refer to ICD website for definition)(32) | Patient-level | = [0.0001,  0.4229,  0.5468,  0.2540,  0.0001,  0.4633,  0.6315,  0.0001,  0.0798,  0.0001,  -0.1023,  0.2328,  0.4569,  0.0001,  -0.0923,  -0.2404,  0.0001,  0.7531,  0.0001,  2.8454,  1.1696,  0.7725,  0.2256,  0.9412,  0.4121,  0.3168,  0.4348,  0.5995,  0.4905,  0.8657,  0.7900] | = [0.0001,  -0.8228,  -0.5327,  -0.6094,  0.0001,  -0.6840  -0.5349,  0.0001,  -0.7336,  0.0001,  -0.7064,  -0.5250,  -0.6761,  0.0001,  0.1349,  -2.0708,  0.0001,  -0.7385,  0.0001,  0.1372,  -0.8852,  -0.5154,  -0.5671,  -0.6998,  -0.7781,  -0.1311,  -0.6141,  -0.8716,  -1.0801,  -0.9450,  -0.6802] |
| Cumulative Charlson comorbidity | Continuous covariates: sum of the binary charlson index | Patient-level | =0.2064 | =0.2887 |
| Cumulative Elixhauser Comorbidity | Continuous covariates: sum of the binary Elixhauser Comorbidity | Patient-level | =-0.5529 | =-0.7735 |
| Cancer type:  cancer_c250pancreatric cancer_c251pancreatric cancer_c252pancreatric  cancer_c253pancreatric cancer_c254pancreatric cancer_c257pancreatric cancer_c258pancreatric  cancer_c259pancreatric | Binary indicator of cancer tumour type. (Please refer to Nation cancer institute for definition of the index)(32) | Patient-level | = [1.4075,  -4.0277,  -5.0587,  -0.7763,  -2.9712,  -1.9126,  -1.8827,  0] | = [0.0626,  0.2530,  0.1875,  -0.0399,  0.43575,  0.1009,  0.2340,  -0.0589] |
| Race:  Black  White  Native Hawaiian/pacific Islander  Others | Binary indicator | Patient-level | =[0,0,0,0] | =[0.0531,  0.1124,  0.2005,  0] |
| Attending physician specialty:  General  Surgical Oncology  Transplant Surgery  Other | Binary indicator | Patient-level | =[0.0001,  -0.0217,  0.4560,  -0.6445] | =[0.0001,  0.2154,  -0.1095,  -0.0453] |
| Length of stays (days) | Continuous covariate | Patient-level | =0.0251 | =0.1413 |

Figure s2: Summary of the various scenarios generated by the plasmode simulation.

| Simulated scenario | Resampled Variables | Beta value to treatment allocation | Beta value to treatment outcome |
| --- | --- | --- | --- |
| Fixed hospital effect on treatment allocation but varying on treatment allocation | Hospital location: urban/rural | Negligible: = 0.002  Low: = 0.2338  Medium: = 0.4675  High: = 0.9350 | Fixed: = -0.8570 |
|  | Hospital yearly surgery volume | Negligible: = 0  Low: = 0.0008  Medium: = 0.0016  High: = 0.0024 | Fixed: = 0.0059 |
|  | Hospital size: beds 500+/ 500> | Negligible: = 0.0009  Low: = 0.0889  Medium: = 0.1778  High: = 0.3556 | Fixed: = 0.3442 |
|  | Hospital region:  Midwest  Northeast  South  West | Negligible: = [0.0057, 0.0021, 0.0022,0]  Low: = [0.5710, 0.2145, 0.2167,0]  Medium: = [1.1419, 0.4290, 0.4333,0.0001]  High: = [2.2838, 0.8580, 0.8666,0] | Fixed: = [0.0990, 0.3626, 0.2016, 0.0001] |
